# SARS-CoV-2 testing of 11,884 healthcare workers at an acute NHS hospital trust in England: a retrospective analysis

**DOI:** 10.1101/2020.12.22.20242362

**Authors:** Aidan T. Hanrath, Ina Schim van der Loeff, Dennis W. Lendrem, Kenneth F. Baker, David A. Price, Peter McDowall, Kiera McDowall, Sue Cook, Peter Towns, Ulrich Schwab, Adam Evans, Jill Dixon, Jennifer Collins, Shirelle Burton-Fanning, David Saunders, Jayne Harwood, Julie Samuel, Matthias L. Schmid, Lucia Pareja-Cebrian, Ewan Hunter, Elizabeth Murphy, Yusri Taha, Brendan A. I. Payne, Christopher J.A. Duncan

**Author notes:** ^*^**Corresponding Author:** Dr Christopher Duncan, Translational and Clinical Research Institute, Newcastle University, UK, NE2 4HH.

## Abstract

Healthcare workers (HCWs) are known to be at increased risk of infection with SARS-CoV-2, although whether these risks are equal across all roles is uncertain. Here we report a retrospective analysis of a large real-world dataset obtained from 10 March to 6 July 2020 in an NHS Foundation Trust in England with 17,126 employees. 3,338 HCWs underwent symptomatic PCR testing (14.4% positive, 2.8% of all staff) and 11,103 HCWs underwent serological testing for SARS-CoV-2 IgG (8.4% positive, 5.5% of all staff). Seropositivity was lower than other hospital settings in England but higher than community estimates. Increased test positivity rates were observed in HCWs from BAME backgrounds and residents in areas of higher social deprivation. A logistic regression model adjusting for these factors showed significant increases in the odds of testing positive in certain occupational groups, most notably domestic services staff, nurses and health-care assistants. PCR testing of symptomatic HCWs appeared to underestimate overall infection levels, probably due to asymptomatic seroconversion. Clinical outcomes were reassuring, with only a small minority of HCWs with COVID-19 requiring hospitalisation (2.3%) or ICU management (0.7%) and with no deaths. Despite a relatively low level of HCW infection compared to other UK cohorts, there were nevertheless important differences in test positivity rates between occupational groups, robust to adjustment for demographic factors such as ethnic background and social deprivation. Quantitative and qualitative studies are needed to better understand the factors contributing to this risk. Robust informatics solutions for HCW exposure data are essential to inform occupational monitoring.

## INTRODUCTION

The pandemic of SARS-CoV-2 serves to highlight the risk posed to healthcare workers (HCWs) by transmissible respiratory pathogens (1-7). As is the case for other highly pathogenic coronaviruses such as severe acute respiratory syndrome (SARS) and Middle East respiratory syndrome (MERS) coronaviruses, SARS-CoV-2 may also be transmitted in healthcare environments (8, 9). Protecting patients and HCWs from nosocomial novel coronavirus-19 disease (COVID-19) is a priority in the control of the SARS-CoV-2 pandemic (1, 10). There are multiple strands to this effort, including environmental controls, use of appropriate personal protective equipment (PPE), as well as rapid testing and the self-isolation at home of SARS-CoV-2 infected HCWs.

Approaches to HCW testing include: (i) PCR testing of those with symptoms (4, 11, 12) or (ii) universal PCR screening (13, 14), recognising that up to 40% of infections may be asymptomatic (15). Each strategy has its limitations and the optimal approach remains to be determined. This decision must balance the risk to HCWs and patients with pragmatic concerns about resource allocation and maintaining safe levels of staffing. Antibody testing adds complementary, albeit retrospective, information about SARS-CoV-2 exposure. Together with PCR testing this provides a resource that can be analysed to inform HCW infection risk.

Recent data suggest that HCWs from certain demographic backgrounds or occupational groups may have different risks of infection (2, 7, 16). To explore this further, we retrospectively analysed a large real-world testing dataset obtained between 10 March and 6 July 2020 in an NHS Foundation Trust in England with 17,126 employees. In this setting, 3,338 HCWs underwent symptomatic PCR testing and 11,103 HCWs underwent antibody testing. The aims of the analysis were: (i) to describe the results of SARS-CoV-2 PCR and antibody testing in this population; (ii) to explore demographic and occupational factors associated with SARS-CoV-2 test positivity, thereby informing the approach to protecting HCWs against COVID-19 in preparation for the next stages of the SARS-CoV-2 pandemic.

## METHODS

### Ethics

As a study of healthcare-associated infections, this was exempt from ethical approval under Section 251 of the NHS Act 2006 and as a study of COVID-19 was also covered by Regulation 3(4) of the Health Service Control of Patient Information Regulations 2002 (March 2020). The study was registered as a clinical service evaluation with approval from the Medical Director. Data extraction and analysis was approved by the Caldicott Guardian (Reference No. 7566).

### Setting

The Newcastle-upon-Tyne Hospitals (NUTH) National Health Service (NHS) Foundation Trust provides secondary care services to a local population of 302,820 (17) and is a tertiary referral centre for the wider North East England and North Cumbria regions. During the period of analysis 17,126 staff were employed across two hospital sites, community sites as well as one offsite non-clinical hub with co-location of administrative, information technology, finance and other support services. NUTH also contains one of two principal contact High Consequence Infectious Diseases (HCID) treatment centres and was the first HCID unit in the UK to manage patients with COVID-19 (18).

### Hospital infection control

From January 2020 there was a focus in the UK on active case identification in people with epidemiological risk of SARS-CoV-2 exposure (contact with a confirmed case or travel to an area with widespread transmission). All suspected or confirmed cases were admitted to HCID units. By March it became clear from hospital admission data that widespread community transmission was occurring. Testing was restricted to hospitalised patients with compatible symptoms. During this period nationwide ‘lockdown’ measures were implemented, including closure of schools, businesses, and travel restrictions for all but essential workers, including NHS workers, on 23 March 2020. Public Health England (PHE) issued regularly updated guidance on personal protective equipment (PPE) for HCWs in NHS hospitals and this guidance was followed in our organisation throughout. Briefly, ‘enhanced’ or ‘level 2’ PPE (FFP3 mask, eye protection (visor), hood, surgical gown, gloves, waterproof apron) was used for contact with all suspected or confirmed patients until 8 March. This was then downgraded to ‘level 1’ PPE (surgical mask, risk-assessed eye protection, apron, gloves) for all patient contacts except those involving aerosol generating procedures (AGPs), which remained at level 2. From 1 April, level 1 PPE was mandated for all care episodes regardless of the patient’s SARS-CoV-2 infection status, except for high-risk clinical areas (such as HDU/ICU) where level 2 PPE was used throughout. From 15 June 2020, surgical facemasks were mandated for all workers in NHS hospitals regardless of patient contact. In NUTH these guidelines were followed and implemented in real time, and PPE was made available to all staff members requiring it. Training was rolled out to all staff members across the Trust with particular attention given to staff members working in environments caring for patients with suspected COVID-19.

### SARS-CoV-2 testing programme

The NUTH staff testing programme has been described elsewhere (19). Briefly, this was jointly developed by the NUTH Occupational Health and Infection Prevention and Control teams. PCR testing of a nasopharyngeal swab was offered to HCWs who were deemed to fulfil the PHE case definition for COVID-19 from 10 March 2020, with a view to early identification of SARS-CoV-2 infected HCWs and to reduce the need for HCWs to self-isolate without knowledge of their infection status. This was in line with the model recommended by NHS England on 12 April 2020. A local modification made by NUTH on 9 April was the inclusion in the case definition of loss of sense of smell (anosmia) and/or taste (ageusia), predating the same change to national guidance on 18 May 2020. HCWs who developed COVID-19 symptoms were advised to immediately self-isolate, contact occupational health by email, and then undergo a nurse administered swab for PCR testing within 3 days (and not greater than 5 days) of the onset of symptoms. Providing that the swab was negative and the HCW considered themselves sufficiently recovered they could return to work. Those who tested positive were advised to remain off work for at least 7 days and until their symptoms resolved (with the exception of a persistent cough or anosmia). As in other NHS settings, PCR testing was undertaken on PHE platforms, initially using the PHE RdRp PCR assay, switching to commercial platforms (Altona Diagnostics from 1 April 2020, with the addition of Roche cobas 6800 from 7 April 2020). In addition, from 29 May 2020, a programme of voluntary testing of SARS-CoV-2 IgG antibody was offered to all NUTH employees. SARS-CoV-2 nucleocapsid IgG testing was undertaken on the Roche platform (Roche Anti-SARS-CoV-2 serology assay).

### Data collection

Data on all PCR and SARS-CoV-2 antibody (Ab) tests undertaken by the regional virology diagnostic laboratory during the period 10 Mar to 6 July 2020 were obtained from a prospectively maintained internal database. In addition, data from the NUTH Electronic Staff Record (ESR) were extracted to obtain demographic information (age, gender, ethnicity, staff role, postcode) of all HCWs employed by NUTH during the same period. Data for certain HCW groups not directly employed by NUTH were unavailable in ESR, therefore these groups were excluded. This included doctors at core and specialty trainee level who are employed by Health Education England North East, and North-East Ambulance Service staff. Data from ESR were matched to virology results data using surname and date of birth, with matching validated by first name, using a script written in Excel (Microsoft). Postcode data were used to obtain data on deprivation index from the Ministry of Housing, Communities and Local Government http://imd-by-postcode.opendatacommunities.org/imd/2019. Staff were assigned to twelve roles based on job title, clinical directorate and specific place of work (Appendix 1). To investigate clinical outcomes of HCWs testing positive for SARS-CoV-2, we cross-referenced testing data with a retrospective database of COVID-19 inpatients managed in NUTH (20), and also searched for additional cases beyond the censor point of this analysis using the electronic inpatient record. Data on hospitalisation, intensive care unit (ICU) admission, ventilation and outcome were collected.

### Data analysis

Measures of central tendency and distribution were calculated using GraphPad Prism version 8.4.3 (GraphPad Software LLC, US). For the initial analysis of demographic factors, ethnicity data were categorised as either white (including white British, white Irish, white other) or black, Asian or any other minority ethnic background (BAME). Deprivation index was categorised into quartiles with the most deprived quartile taken as the reference group. Contingency tables and Chi^2^ (χ^2^) tests were used to compare positivity rates between groups. Differences in positivity rates between staff roles were estimated using a multiple logistic regression model to adjust for the effects of age, ethnicity (BAME), gender, and deprivation (deciles were used for this analysis). Regression modelling was performed using the SAS JMP Pro Statistical Visualization Software (JMP, UK). A dummy variable (phase) was created to assess for any interaction between staff roles and the proportions of HCWs presenting for antibody testing with and without a prior history of presentation for PCR testing. In addition, the robustness of the staff roles effect was examined by stepping candidate covariables in and out of the logistic regression supplemented by generalized linear regression. While the Ab positivity rates were higher for those presenting with a prior history of PCR testing, there was no statistically significant interaction between Staff Roles and Phase (p=0.6963). The interaction term was dropped, and the odds ratios and 95% confidence intervals constructed for the comparison of each of the Staff Roles relative to the minimal exposure group (Administrative and Managerial).

## RESULTS

### PCR testing

From 10 March to 6 July 2020, NUTH laboratories processed and provided SARS-CoV-2 PCR results on 44,781 combined nose/throat swabs. During this period, 3,721 PCR tests were undertaken on 3,338 HCWs who had contacted the symptomatic testing programme (representing 19.5% of all NUTH employees). The median (IQR) turnaround time from samples arriving in the laboratory to a result being available was 7.8 (6.5 – 10.5) hours. In total 481/3,338 symptomatic HCWs tested positive for SARS-CoV-2 by PCR (14.4% [95% CI 13.3-15.6%] of those tested; 2.8% [2.6-3.1%] of all HCWs in the organisation).

### PCR positivity rates varied over time

The number of HCWs presenting for testing and the rate of positive tests fluctuated during the study period, corresponding to the dynamics of SARS-CoV-2 transmission in the region (**Figure 1**). The number of tests performed per day ranged from three to 169 (**Figure 2A**). Most positive PCR tests (390/481, 80%) were returned in the four weeks between 23 March and 19 April, when around half of all PCR tests were done (1959/3721 [52.6%]). In this period the seven day average per-test positivity rate peaked at 23.9%, before decreasing and becoming more variable as the number of tests performed on symptomatic staff reduced (**Figure 2B**). Per-test positivity rates (seven-day average) in the last 4 weeks of the testing period were 0.8%, 2.6%, 0.0% and 0.0%, when there were only three positive tests in total.

**Figure 1.**
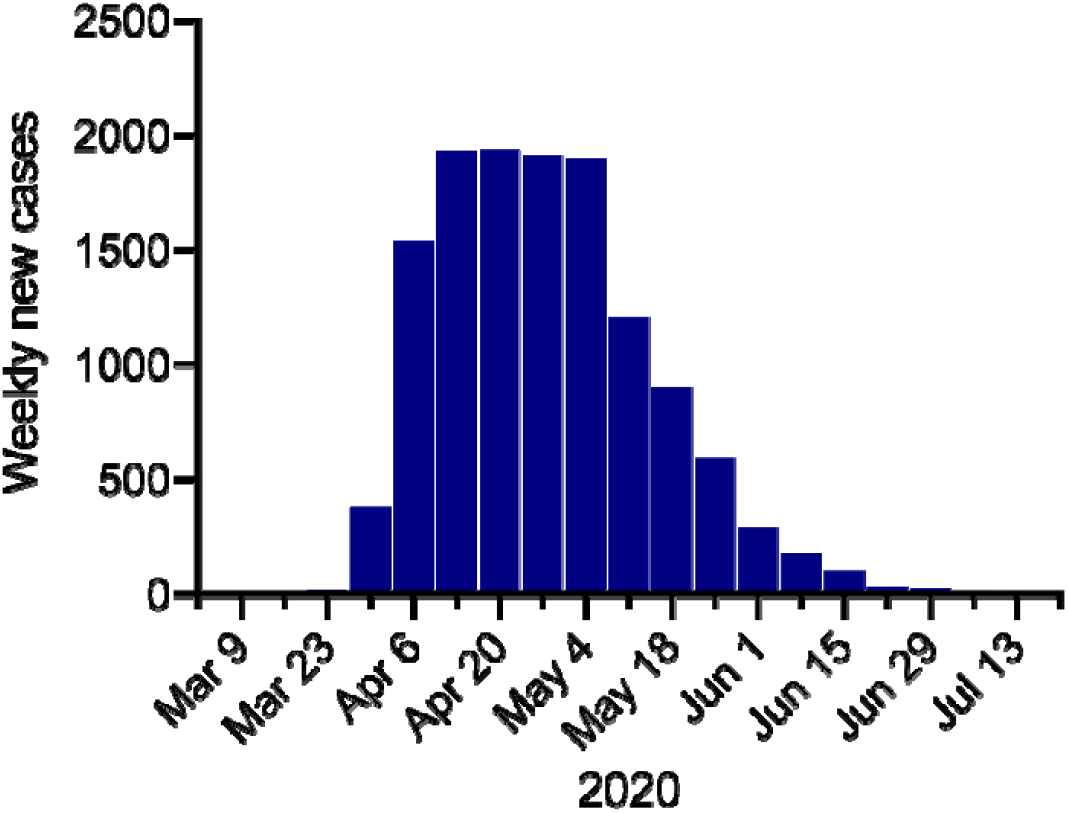
Cases of COVID-19 in the North East England region during the study period.

**Figure 2.**
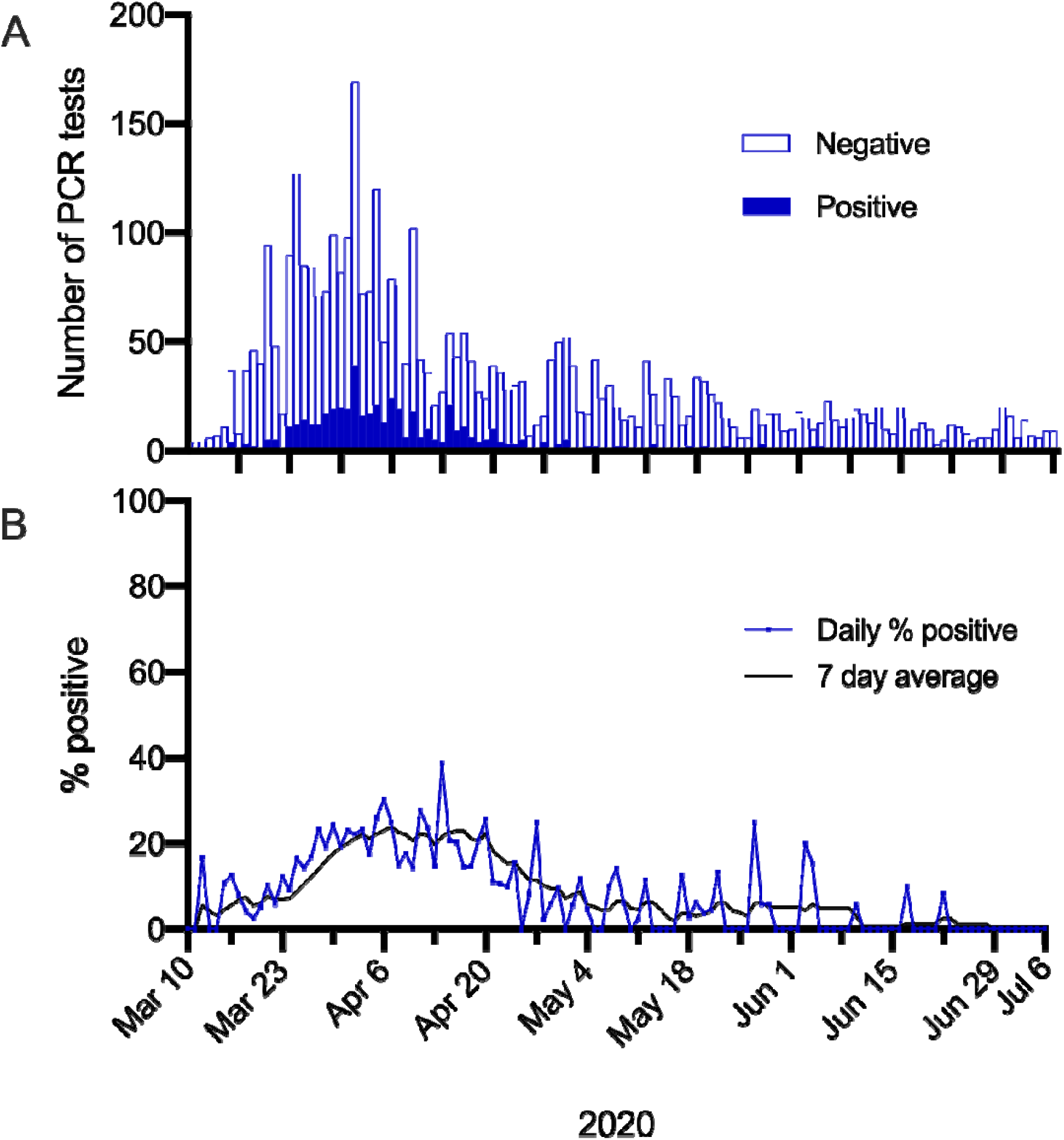
Changes in PCR test positivity over time. (A) Total SARS-CoV-2 PCR tests in HCWs. Filled bars = positive tests. Open bars = negative tests. (B) Per test positivity (%), displayed as daily (blue line) or 7 day average (black line).

### COVID-19 clinical outcomes

To investigate clinical outcomes of HCWs symptomatically infected with SARS-CoV-2, we cross-referenced testing data with a separate database of COVID-19 inpatients managed in NUTH (20), and also searched hospital electronic patient records of PCR positive HCWs for additional cases beyond the censor date of this prior analysis. Seventeen of 481 (3.5%) HCWs testing positive were assessed in secondary care, and 10 (2.1%, 0.06% of all staff) required hospital admission. The median (IQR) [range] length of stay was 5 (3-8.5) [1-12] days. Three PCR-positive HCWs had severe disease on admission defined according to World Health Organisation (WHO) criteria (oxygen saturations <90% without supplemental oxygen and/or respiratory rate > 30 breaths/min) and three (0.6%) were managed in critical care, two with non-invasive pressure support. No patients were intubated or required extracorporeal membrane oxygenation (ECMO). All survived to hospital discharge.

### Antibody testing

To complement the PCR analysis, data were analysed from a voluntary seroprevalence survey which was available to all HCWs irrespective of role and/or prior PCR testing and widely advertised in the organisation, from the 29 May. 11,103 of 17,126 HCWs (64.8%) came forward for antibody testing, including 2,557 HCWs who had previously undergone PCR testing (**Figure 3**). SARS-CoV-2 IgG was detected in 937/11,103 (8.4%) HCWs (5.5% of all staff). A gradient of seropositivity was observed, from 380/409 (92.9% [95% CI 90.0-95.0%]) of those testing positive by PCR, to 161/2,148 (7.5% [6.5-8.7%]) of those testing negative by PCR, and 396/8,546 (4.6% [4.2-5.1%]) of those who had not had a PCR test (P<0.001, χ^2^ test).

**Figure 3.**
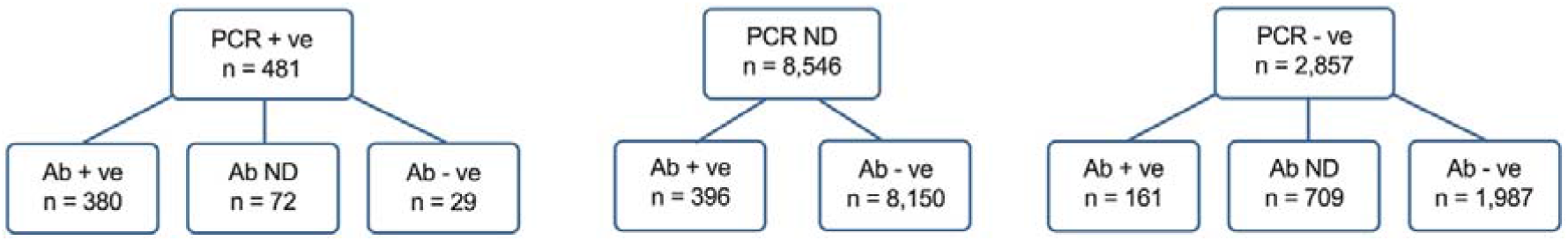
Schematic demonstrating SARS-CoV-2 tests done in the study population. ND = not done.

### Demographic factors associated with seropositivity

There was no difference in the median (IQR) age of HCWs with detectable or undetectable SARS-CoV-2 IgG antibody (median 43 [IQR: 30-54] and 43 [32-53] years respectively, t test). 734/8,549 (8.6% [95% CI 8.0-9.2]) females were seropositive compared to 150/2,037 (7.4% [6.3-8.6]) males (χ^2^ test p = 0.073). Seropositivity in HCWs of white ethnicity was 774/9,500 (8.1% [95% CI 7.6-8.7] percent), compared to 95/894 (10.6% [8.8-12.8]) in those from BAME backgrounds (χ^2^ test p = 0.011). Comparing deprivation data, seropositivity was noted in 301/2,926 (10.3% [95% CI 9.2-11.4]) of HCWs from the most deprived quartile, compared to 575/7,571 (7.6% [7.0-8.2]) of the less deprived three quartiles (χ^2^test p < 0.001).

### Association of HCW role with SARS-CoV-2 infection

To explore associations between occupational role and the proportion of positive tests (defined as individuals with a positive test by PCR and/or antibody as a percentage of all those tested), HCWs were grouped into 12 categories based on roles recorded in ESR (as discussed in Appendix 1). Logistic regression analysis was performed adjusting for the demographic factors described above (**Table 1**). The administrative and managerial, non-patient facing group was used as the comparator for this analysis based on the fact that their role does not require close contact with patients or the hospital environment and that many of these staff work in an off-site location separate from the hospital sites.

### Antibody testing and PCR testing

Following adjustment for age, sex, ethnicity and deprivation decile there was strong statistical evidence of differences in positivity rates across staff roles for both antibody and PCR testing (p<0.0001). Most notably, the odds of having a positive antibody test were greater for domestic services staff, healthcare assistants (HCA) and nurses, in addition to estates and catering and patient-facing clerical workers (**Figure 4A**). A similar pattern was observed for PCR testing with the odds of testing positive also being greater for domestic Services staff, HCA and nurses (**Figure 4B**). Adjusted odds ratios and 95% confidence intervals for antibody and PCR positivity for each of the roles relative to administrative and managerial workers (the reference group) are shown in **Figure 4A-B**. For reference, the raw data are included in **Supplementary Table 1**.

**Figure 4.**
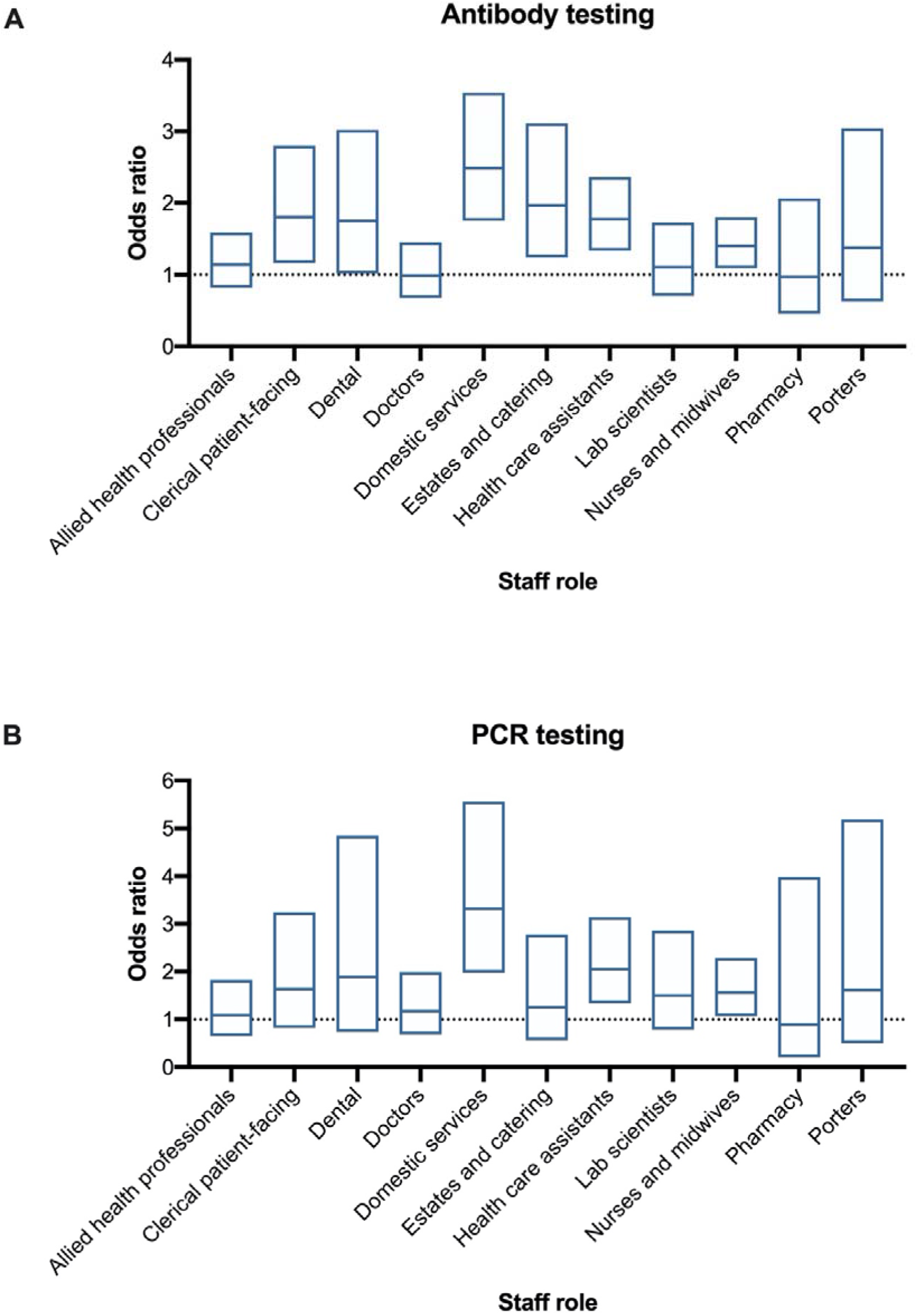
Differential test positivity among HCW groups. (A) Adjusted odds ratio of a positive Ab test by staff category. (B) Adjusted odds ratio of a positive PCR test by staff category. Adjusted odds ratio (central line) and 95% confidence interval (box) calculated by logistic regression as described in text (compared to non-patient facing administrative and managerial workers).

## DISCUSSION

The data we report here span the first wave of the SARS-CoV-2 epidemic in England and represent among the largest combined molecular and serological testing datasets in a HCW population. Nearly one in five employees in this large organisation presented for PCR testing during the study period and 14.4% percent of those tested (2.8% of the workforce) had symptomatic SARS-CoV-2 infection detected by PCR. Over two thirds of the total workforce (over 10,000 HCWs) underwent antibody testing. 8.4% of those tested (5.5% of the workforce) were seropositive. This compares to seroprevalence estimates of 6.0% for England and 5.0% for the North East of England around the same period (21) and is consistent with increased exposure in HCWs.

These positivity rates are considerably lower than rates among HCWs in some areas of England, such as London (3), Birmingham (7), and in other parts of the North East (12), although are similar to other regions such as Oxford (2) and Cambridge (14). Factors determining the regional variation in HCW infection rates are unknown, although a relationship with the burden of inpatient cases is apparent (2, 3, 7). It was not possible to draw direct comparisons with community PCR positivity rates, due to the absence of community testing during this period in England. However, community transmission can be inferred from hospital admission data. We note that PCR-confirmed cases among HCWs fell during the study period, in parallel with the decline in community and hospital cases. This occurred despite the fact that most HCWs continued to commute to work and mix in the hospital environment. Similar observations were made at another NHS site (14, 22). No shortages of PPE were reported in our organisation. This along with HCW training in donning and doffing PPE might have helped to reduce seroprevalence amongst our staff. These data suggest that the risk of sustained HCW-to-HCW transmission of SARS-CoV-2 can be mitigated in hospital environments (22), despite the recognised challenge of physical distancing in these and other healthcare settings (23).

In our analysis, baseline factors associated with seroconversion included being from black, Asian and minority ethnic (BAME) backgrounds, and living in areas of greater social deprivation, consistent with published data from both HCWs (2, 7) and the general population (21). Our analysis makes the important additional contribution of showing that test positivity rates differ by occupational role, including after adjustment for contributing demographic factors. These occupational differences cut across clinical and non-clinical roles. Compared with the comparator group of administrative and management workers, nurses and midwives as well as healthcare assistants and dental hospital workers were more likely to test positive whereas doctors or allied health professionals did not, suggesting factors beyond patient contact may be involved.

Other interesting observations also emerged from the analysis. Among non-clinical HCWs exposed to the hospital environment, domestic services and estates/catering workers were more likely to test positive, whereas laboratory workers handling potentially infectious specimens were not. Administrative staff working in the hospital environment (such as receptionists and ward clerks) had higher positivity rates than those outside it. The underlying reasons for differing rates among occupational groups are not known. An important limitation to the analysis was that details on individuals’ contact with cases of COVID-19, either at home or in the workplace, was not collected routinely. This was in part due to how the HCW testing programme was developed, i.e. rapidly and under conditions of extremely high demand. In parallel there was also an unprecedented redeployment of HCWs to COVID-19 areas for clinical service provision throughout the organisation. This change in activity was not captured in the ESR. The value of collecting this information was demonstrated recently in another UK study where similar differences in seroprevalence by occupation were noted, including increased seroprevalence rates in domestic services, porters, nurses and estates and catering staff, although only increased rates among domestic services staff and porters (as a combined group) remained significant after adjustment for exposure to COVID-19 (2). Other studies in the UK have not reported rates according to individual occupational roles (3, 7, 11), although did highlight an increased risk among ‘housekeeping’ workers (7) - equivalent to domestic services workers in our dataset. Thus there is an emerging picture of higher seroprevalence rates among domestic services workers as well as those HCWs from BAME backgrounds (2, 7). Whilst the underlying reasons for this are likely to be multifactorial and to include economic and social factors, enhanced surveillance and/or targeted infection control measures are a priority in these groups.

So too are further studies to understand the relative contribution of risks. It is worth noting that within NUTH, domestic services staff used level 1 PPE from 8 March onwards. Our data also provide a signal of heightened risk in other occupational groups, notwithstanding the limitations described above. Analysis of the reproducibility of these observations in other datasets is justified. For example, some studies have shown nursing staff to be at increased risk of acquiring both SARS-CoV-2 (16, 24) and SARS-CoV (25), while others have not (26, 27). Duration of patient contact (16) and incorrect use of PPE (28-32) have also been cited as potential contributing factors in SARS-CoV-2 acquisition in health care settings. Whilst occupational risk is often the focus (3, 6, 13, 16), studies continue to highlight the contribution of community acquisition (2, 5, 33, 34). Until the underlying reasons for differential rates of positivity between occupational groups are established it will be important to continue to monitor infection rates in future waves of SARS-CoV-2 transmission to assess whether current risk mitigation strategies are sufficient. Our findings also highlight the urgent need for robust informatics solutions to allow for routine collection of exposure data at an organisational level.

This study has additional limitations. Data were collected retrospectively, thus are more prone to bias. Testing relied on HCWs presenting with symptoms or coming forward for antibody testing, therefore positives may have been missed in both cases, or alternatively this strategy may have selected for those at greater risk of testing positive. It was also not possible to account for a minority of HCWs who were shielding, and thus at much lower exposure risk, although this issue is likely to be shared across all occupational groups. Finally, small numbers made it necessary to pool some groups for analysis, resulting in relatively arbitrary staff categories (such as estates and catering or dental hospital workers).

A strength of this dataset, compared to other published studies, is the opportunity it provides to compare results of PCR and subsequent antibody testing in over 2,500 individual HCWs. Seropositivity was 93% in those with prior PCR-confirmed infection. These data are informative as there are few studies of seroconversion rates in HCWs or in people with mild COVID-19 confirmed by prior PCR testing. The results suggest that most patients with mild but symptomatic COVID-19 seroconvert, albeit with a notable minority (7%) who do not. Whether these individuals mount a T-cell response to SARS-CoV-2 is an open question. It is worth noting that in all cases antibody positivity was documented at a time after the positive PCR test, i.e. no PCR-confirmed re-infections occurred. Our data suggest that the ELISA assay is a broadly acceptable surrogate for SARS-CoV-2 exposure in studies of non-hospitalised populations. The observation that seropositivity was higher in those with a negative PCR test than those who had not undergone prior PCR testing is interesting and has been reported elsewhere (27). This is possibly explained by false negative SARS-CoV-2 PCR testing, which can arise through a number of practical (e.g. sampling technique) and methodological issues (e.g. assay design) (35). In mitigation, HCWs tasked with taking swabs underwent extensive training, only pooled nose and throat swabs were taken, and the most sensitive laboratory platforms were used once available.

The symptom-based testing approach we employed appears to have underestimated total HCW infections. The observation that around 4.6% of HCWs who did not present for symptomatic PCR testing were seropositive suggests that a considerable proportion of HCWs either experienced asymptomatic or pauci-symptomatic infection, or that they did not present for PCR testing despite experiencing symptoms. In support of the former hypothesis, a recent meta-analysis has indicated that between 4 and 41% of SARS-CoV-2 infections are asymptomatic (15). A large proportion of cases may be missed by a symptom-based testing approach, consistent with our observations. Recent data in HCWs have confirmed that asymptomatic SARS-CoV-2 infection does occur (2, 3, 7, 13, 14) and this is central to the argument for asymptomatic screening (1). This is a reasonable approach in low incidence settings. However important uncertainties to be balanced against asymptomatic HCW screening are the extent to which asymptomatic HCWs transmit SARS-CoV-2 (15), alongside more pragmatic considerations such as how frequently to screen and how to deal with the issue of prolonged asymptomatic shedding of SARS-CoV-2 RNA, which occurs in between a quarter (2) and a half (36) of HCWs, but is not thought to necessarily represent infectious virus (13, 37). Roll out of asymptomatic testing in healthcare settings is anticipated.

Despite an increased risk of SARS-CoV-2 infection, cumulative mortality rates appear lower in HCWs than in the general UK population (38). Our data demonstrate reassuringly low rates of both hospitalisation and need for critical care. This may be due to the relative absence of risk factors for mortality in this population such as advanced age and comorbidities (20), coupled with earlier diagnosis and access to treatment. This pattern has also been reported in China (24) and the US (39).

In summary, the data reported here demonstrate that despite a relatively low level of infection compared to other UK HCW cohorts, there was an important differential risk of infection between occupational groups, robust to adjustment for other demographic factors such as BAME background and social deprivation. This finding adds to the growing evidence of differential risks among HCWs. In order to better understand the factors contributing to these risks, prospective quantitative and qualitative studies are a priority. In addition, robust informatics solutions to facilitate the routine collection of ‘real world’ clinical data on HCW exposure and testing within the NHS are critical to inform risk assessment and monitoring.

## Data Availability

Please refer requests to corresponding author. Data is protected by NHS Caldicott guidelines.

## Acknowledgements

The authors acknowledge the enormous contribution of the great many nursing, laboratory, administrative, IT and managerial colleagues within NUTH who did the work to generate these data and to the staff of NUTH who came forward for testing. CJAD is funded by the Wellcome Trust (211153/Z/18/Z), the UKRI UK-Coronavirus Immunology Consortium, and the Barbour Foundation. AH is funded by the British Medical Association. KFB is funded by a National Institute for Health Research (NIHR) Clinical Lectureship (CL-2017-01-004). BAIP is funded by the Wellcome Trust (109975/Z/15/Z, 203105/Z/16/Z). The views expressed are those of the authors and not necessarily those of NUTH, the NHS, the NIHR or the Department of Health and Social Care.

## References

1. Black JRM, Bailey C, Przewrocka J, Dijkstra KK, Swanton C. COVID-19: the case for health-care worker screening to prevent hospital transmission. Lancet. 2020;395(10234):1418–20.

2. Eyre DW, Lumley SF, O’Donnell D, Campbell M, Sims E, Lawson E, et al. Differential occupational risks to healthcare workers from SARS-CoV-2 observed during a prospective observational study. Elife. 2020;9.

3. Houlihan CF, Vora N, Byrne T, Lewer D, Kelly G, Heaney J, et al. Pandemic peak SARS-CoV-2 infection and seroconversion rates in London frontline health-care workers. Lancet. 2020;396(10246):e6–e7.

4. Hunter E, Price DA, Murphy E, van der Loeff IS, Baker KF, Lendrem D, et al. First experience of COVID-19 screening of health-care workers in England. Lancet. 2020;395(10234):e77–e8.

5. Muhi S, Irving LB, Buising KL. COVID-19 in Australian health care workers: early experience of the Royal Melbourne Hospital emphasises the importance of community acquisition. Med J Aust. 2020;213(1):44–e1.

6. Nguyen LH, Drew DA, Graham MS, Joshi AD, Guo CG, Ma W, et al. Risk of COVID-19 among front-line health-care workers and the general community: a prospective cohort study. Lancet Public Health. 2020;5(9):e475–e83.

7. Shields A, Faustini SE, Perez-Toledo M, Jossi S, Aldera E, Allen JD, et al. SARS-CoV-2 seroprevalence and asymptomatic viral carriage in healthcare workers: a cross-sectional study. Thorax. 2020.

8. Santarpia JL, Rivera DN, Herrera VL, Morwitzer MJ, Creager HM, Santarpia GW, et al. Aerosol and surface contamination of SARS-CoV-2 observed in quarantine and isolation care. Sci Rep. 2020;10(1):12732.

9. Ye G, Lin H, Chen S, Wang S, Zeng Z, Wang W, et al. Environmental contamination of SARS-CoV-2 in healthcare premises. J Infect. 2020;81(2):e1–e5.

10. Chirico F, Nucera G, Magnavita N. COVID-19: Protecting Healthcare Workers is a priority. Infect Control Hosp Epidemiol. 2020;41(9):1117.

11. Keeley AJ, Evans C, Colton H, Ankcorn M, Cope A, State A, et al. Roll-out of SARS-CoV-2 testing for healthcare workers at a large NHS Foundation Trust in the United Kingdom, March 2020. Euro Surveill. 2020;25(14).

12. Leeds JS, Raviprakash V, Jacques T, Scanlon N, Cundall J, Leeds CM. Risk factors for detection of SARS-CoV-2 in healthcare workers during April 2020 in a UK hospital testing programme. EClinicalMedicine. 2020;26:100513.

13. Brown CS, Clare K, Chand M, Andrews J, Auckland C, Beshir S, et al. Snapshot PCR surveillance for SARS-CoV-2 in hospital staff in England. J Infect. 2020;81(3):427–34.

14. Rivett L, Sridhar S, Sparkes D, Routledge M, Jones NK, Forrest S, et al. Screening of healthcare workers for SARS-CoV-2 highlights the role of asymptomatic carriage in COVID-19 transmission. Elife. 2020;9.

15. Byambasuren O, Cardona M, Bell K, Clark J, McLaws M, Glasziou P. Estimating the extent of asymptomatic COVID-19 and its potential for community transmission: systematic review and meta-analysis. 2020.

16. Barrett ES, Horton DB, Roy J, Gennaro ML, Brooks A, Tischfield J, et al. Prevalence of SARS-CoV-2 infection in previously undiagnosed health care workers at the onset of the U.S. COVID-19 epidemic. medRxiv. 2020.

17. Population Estimates for the UK, England and Wales, Scotland and Northern Ireland: Mid-2019, using April 2020 local authority district codes. Office for National Statistics (2020). https://www.ons.gov.uk/peoplepopulationandcommunity/populationandmigration/populationestimates/datasets/populationestimatesforukenglandandwalesscotlandandnorthernireland [accessed online 17 June 2020].

18. Lillie PJ, Samson A, Li A, Adams K, Capstick R, Barlow GD, et al. Novel coronavirus disease (Covid-19): The first two patients in the UK with person to person transmission. The Journal of infection. 2020;80(5):578–606.

19. Matthewson J, Tiplady A, Gerakios F, Foley A, Murphy E. Implementation and analysis of a telephone support service during COVID-19. Occupational Medicine. 2020.

20. Baker KF, Hanrath AT, Schim van der Loeff I, Tee SA, Capstick R, Marchitelli G, et al. COVID-19 management in a UK NHS Foundation Trust with a High Consequence Infectious Diseases centre: a detailed descriptive analysis. medRxiv. 2020:2020.05.14.20100834.

21. Ward H, Atchison CJ, Whitaker M, Ainslie KEC, Elliot J, Okell LC, et al. Antibody prevalence for SARS-CoV-2 in England following first peak of the pandemic: REACT2 study in 100,000 adults. 2020.

22. Jones NK, Rivett L, Sparkes D, Forrest S, Sridhar S, Young J, et al. Effective control of SARS-CoV-2 transmission between healthcare workers during a period of diminished community prevalence of COVID-19. Elife. 2020;9.

23. Arora VM, Chivu M, Schram A, Meltzer D. Implementing Physical Distancing in the Hospital: A Key Strategy to Prevent Nosocomial Transmission of COVID-19. Journal of hospital medicine. 2020;15(5):290–1.

24. Zheng L, Wang X, Zhou C, Liu Q, Li S, Sun Q, et al. Analysis of the infection status of the health care workers in Wuhan during the COVID-19 outbreak: A cross-sectional study. Clin Infect Dis. 2020.

25. Ip M, Chan PK, Lee N, Wu A, Ng TK, Chan L, et al. Seroprevalence of antibody to severe acute respiratory syndrome (SARS)-associated coronavirus among health care workers in SARS and non-SARS medical wards. Clin Infect Dis. 2004;38(12):e116–8.

26. Schmidt SB, Gruter L, Boltzmann M, Rollnik JD. Prevalence of serum IgG antibodies against SARS-CoV-2 among clinic staff. PLoS One. 2020;15(6):e0235417.

27. Stubblefield WB, Talbot HK, Feldstein L, Tenforde MW, Rasheed MAU, Mills L, et al. Seroprevalence of SARS-CoV-2 Among Frontline Healthcare Personnel During the First Month of Caring for COVID-19 Patients - Nashville, Tennessee. Clin Infect Dis. 2020.

28. Bays DJ, Nguyen MH, Cohen SH, Waldman S, Martin CS, Thompson GR, et al. Investigation of Nosocomial SARS-CoV-2 Transmission from Two Patients to Health Care Workers Identifies Close Contact but not Airborne Transmission Events. Infect Control Hosp Epidemiol. 2020:1–22.

29. Guo X, Wang J, Hu D, Wu L, Gu L, Wang Y, et al. Survey of COVID-19 Disease Among Orthopaedic Surgeons in Wuhan, People’s Republic of China. J Bone Joint Surg Am. 2020;102(10):847–54.

30. Maltezou HC, Dedoukou X, Tseroni M, Tsonou E, Raftopoulos V, Papadima K, et al. SARS-CoV-2 infection in healthcare personnel with high-risk occupational exposure: evaluation of seven-day exclusion from work policy. Clin Infect Dis. 2020.

31. Nienhaus A, Hod R. COVID-19 among Health Workers in Germany and Malaysia. Int J Environ Res Public Health. 2020;17(13).

32. Paderno A, Fior M, Berretti G, Schreiber A, Grammatica A, Mattavelli D, et al. SARS-CoV-2 Infection in Health Care Workers: Cross-sectional Analysis of an Otolaryngology Unit. Otolaryngol Head Neck Surg. 2020:194599820932162.

33. Quattrone F, Vabanesi M, Borghini A, De Vito G, Emdin M, Passino C. The value of hospital personnel serological screening in an integrated COVID-19 infection prevention and control strategy. Infect Control Hosp Epidemiol. 2020:1–2.

34. Steensels D, Oris E, Coninx L, Nuyens D, Delforge ML, Vermeersch P, et al. Hospital-Wide SARS-CoV-2 Antibody Screening in 3056 Staff in a Tertiary Center in Belgium. JAMA. 2020.

35. Woloshin S, Patel N, Kesselheim AS. False Negative Tests for SARS-CoV-2 Infection - Challenges and Implications. N Engl J Med. 2020;383(6):e38.

36. Schwierzeck V, Correa-Martinez CL, Schneider KN, Mellmann A, Hennies MT, Hafezi W, et al. SARS-CoV-2 in the Employees of a Large University Hospital. Dtsch Arztebl Int. 2020;117(19):344–5.

37. Wolfel R, Corman VM, Guggemos W, Seilmaier M, Zange S, Muller MA, et al. Virological assessment of hospitalized patients with COVID-2019. Nature. 2020;581(7809):465–9.

38. Levene LS, Coles B, Davies MJ, Hanif W, Zaccardi F, Khunti K. COVID-19 cumulative mortality rates for frontline healthcare staff in England. Br J Gen Pract. 2020;70(696):327–8.

39. Team CC-R. Characteristics of Health Care Personnel with COVID-19 - United States, February 12-April 9, 2020. MMWR Morb Mortal Wkly Rep. 2020;69(15):477–81.

