## Supplemental Table 1 for "SARS-CoV-2 testing of 11,884 healthcare workers at an acute NHS hospital trust in England: a retrospective analysis"

**Supplementary Table 1**. Proportion of positive tests among staff groups. Odds ratio and 95% confidence intervals calculated by logistic regression as described in text (*in relation to the comparator group of non-patient facing administrative and managerial workers). Dental hospital staff included a combination of professional groups and were pooled due to small numbers.

|  | Number of staff | PCR positive/tested (%) | Adjusted odds ratio (95% CI) | Ab positive/tested (%) | Adjusted odds ratio (95% CI) |
| --- | --- | --- | --- | --- | --- |
| Admin and managerial (non patient-facing)* | 2,530 | 41/399 (10.28%) | - | 101/1,726 (5.85%) | - |
| Allied health professionals | 1,681 | 30/305 (9.84%) | 1.09 (0.65-1.83) | 73/1,102 (6.62%) | 1.14 (0.82-1.59) |
| Clerical patient-facing | 415 | 14/86 (16.28%) | 1.63 (0.82-3.24) | 33/315 (10.48%) | 1.8 (1.16-2.8) |
| Dental hospital staff | 358 | 7/41 (17.07%) | 1.89 (0.74-4.85) | 20/244 (8.2%) | 1.75 (1.02-3.02) |
| Doctors | 1,746 | 44/361 (12.19%) | 1.17 (0.69-1.99) | 63/899 (7.01%) | 0.99 (0.67-1.45) |
| Domestic services | 813 | 37/137 (27.01%) | 3.32 (1.98-5.56) | 68/515 (13.2%) | 2.49 (1.75-3.54) |
| Estates and catering | 517 | 10/72 (13.89%) | 1.25 (0.56-2.78) | 29/340 (8.53%) | 1.97 (1.24-3.12) |
| Health care assistants | 2,351 | 89/481 (18.5%) | 2.05 (1.34-3.14) | 164/1,434 (11.44%) | 1.78 (1.33-2.36) |
| Lab scientists | 643 | 15/122 (12.3%) | 1.5 (0.78-2.86) | 29/468 (6.2%) | 1.1 (0.71-1.73) |
| Nurses and midwives | 5,536 | 188/1,281 (14.68%) | 1.56 (1.07-2.29) | 337/3,749 (8.99%) | 1.4 (1.09-1.8) |
| Pharmacy | 304 | 2/28 (7.14%) | 0.89 (0.2-3.98) | 9/189 (4.76%) | 0.97 (0.45-2.06) |
| Porters | 232 | 4/25 (16%) | 1.61 (0.5-5.19) | 11/122 (9.02%) | 1.38 (0.62-3.05) |
